# Triglyceride Polygenic Score Identifies Differential Bleeding and Cardiovascular Risk with Aspirin in Primary Prevention

**DOI:** 10.64898/2026.02.19.26346656

**Authors:** Peter D. Fransquet, Chenglong Yu, Cammie Tran, Sultana Monira Hussain, Chad A. Bousman, Mark R. Nelson, Andrew M. Tonkin, John J. McNeil, Paul Lacaze

## Abstract

**Aims:** Low-dose aspirin is no longer routinely recommended for primary prevention in older adults because bleeding risks outweigh cardiovascular benefits. We aimed to investigate whether polygenic scores (PGSs) could modify the effects of aspirin on major bleeding and major adverse cardiovascular events (MACE) in a trial of older individuals.

**Methods:** We conducted *post-hoc* genetic analysis of the Aspirin in Reducing Events in the Elderly (ASPREE) randomized, placebo-controlled trial in Australia and the United States. Participants aged ≥70 years (≥65 years for U.S. minorities) without cardiovascular disease, dementia, or physical disability were randomized to 100 mg daily aspirin or placebo. Among those with high-quality genotyping data (n=13,571; median follow-up 4.6 years), we tested 572 cardiovascular- and hematologic-related PGSs for interaction with aspirin using Cox proportional hazards models, applying Bonferroni correction.

**Results:** A triglyceride-related PGS (PGS003144) modified aspirin’s effect on major bleeding (interaction P=5.9×10^−5^; Bonferroni-adjusted P=0.034). In the lowest PGS quintile, aspirin increased major bleeding compared with placebo (hazard ratio [HR] 2.28; 95% CI 1.45–3.58) without reducing MACE (HR 1.04; 95% CI 0.67–1.62). In contrast, in the highest quintile, aspirin was associated with lower risks of major bleeding (HR 0.62; 95% CI 0.38–0.97) and MACE (HR 0.66; 95% CI 0.44–0.99). Baseline measured triglyceride levels demonstrated a similar pattern of effect modification.

**Conclusion:** A triglyceride-related PGS identifies older adults with divergent bleeding and cardiovascular responses to aspirin, supporting the potential role of genetically-informed strategies for primary cardiovascular prevention.

**Lay summary:** This study shows that genetic differences related to triglyceride levels may help identify older adults who are more likely to be harmed or to benefit from taking aspirin to prevent heart disease.

- In older adults with certain genetic profiles linked to triglycerides, aspirin increased the risk of serious bleeding without reducing heart attacks or strokes, while in others it was associated with lower risks of both bleeding and cardiovascular events.
- Using genetic information alongside traditional risk factors could help tailor aspirin use for primary prevention, avoiding unnecessary harm while identifying those most likely to benefit.

**Graphical Abstract:** Figure, Graphical Abstract:
Genetic stratification of aspirin benefit and harm using a triglyceride polygenic score.
Screening of 572 polygenic scores in the ASPREE trial identified a triglyceride-related PGS that modified aspirin-associated bleeding and cardiovascular risk. Aspirin increased bleeding risk in the lowest PGS quintile but reduced major bleeding and MACE in the highest quintile.
**Abbreviations:** PGS, polygenic score; GI, gastrointestinal; IC, intracranial; MACE, major adverse cardiovascular events.

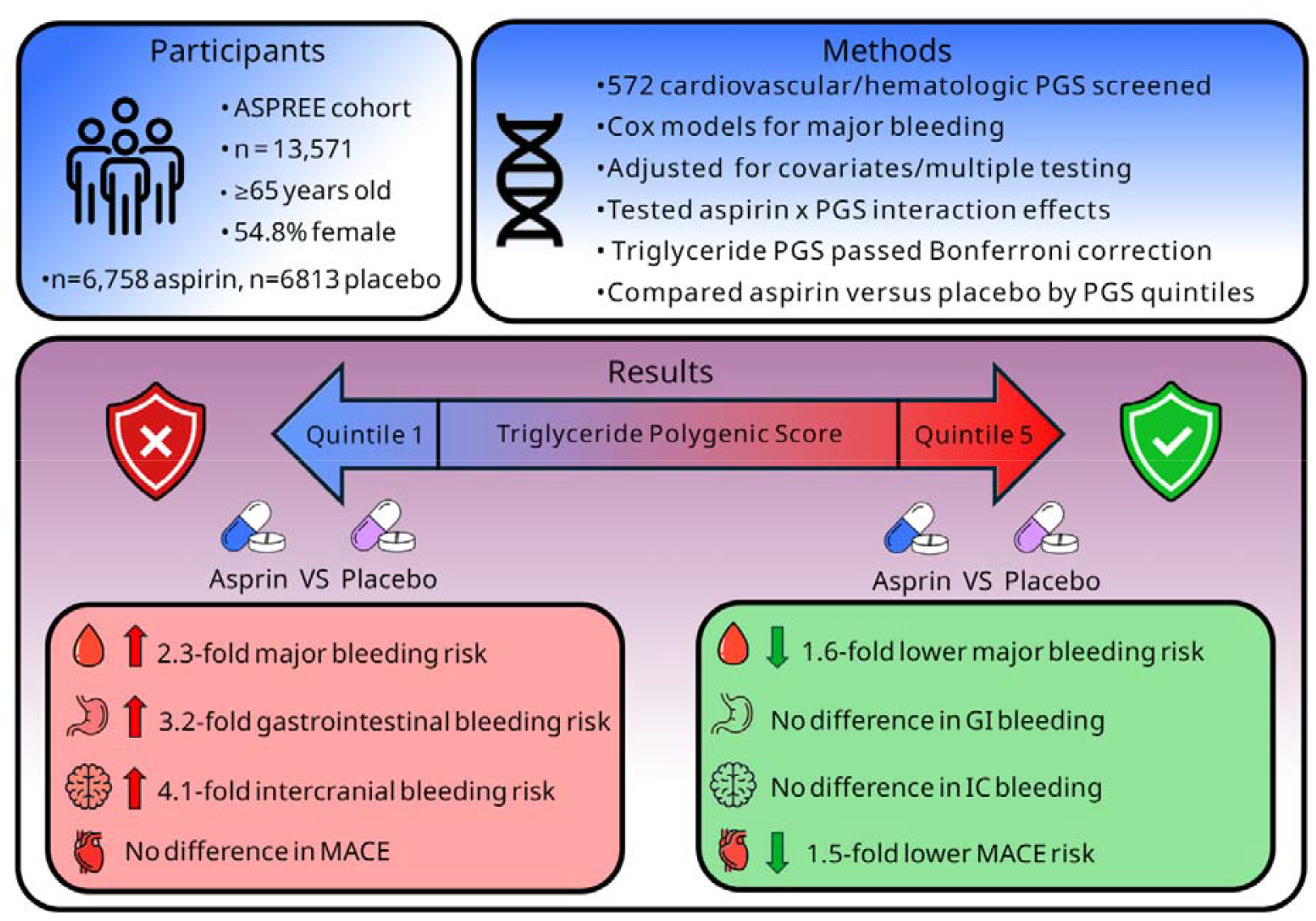

## INTRODUCTION

Low-dose aspirin has historically been widely used for the primary prevention of atherothrombotic cardiovascular disease events in older people. But recent evidence has tempered the use of aspirin in this setting, due to increased bleeding risks amidst lack of vascular benefits ^1-4^. Three large, randomized trials (ARRIVE, ASCEND, and ASPREE) ^3,5,6^ all reported net unfavorable benefit:harm ratios for aspirin, due to bleeding risks. None of these trials, however, accounted for the potential underlying interindividual or genetic difference in aspirin responses, that may influence risk of bleeding or cardiovascular events. Identifying factors that may differentiate or stratify individuals regarding aspirin response could provide a pathway towards more personalized therapeutic use. Consistent with this concept, a large benefit:harm analysis previously demonstrated that a small but clinically meaningful proportion of individuals without established cardiovascular disease may derive net benefit from aspirin when cardiovascular and bleeding risks are considered jointly, highlighting the potential value of individualized approaches to aspirin prescribing ^7^. Further, our previous work suggested that subgroups of the ASPREE trial population with specific lipoprotein(a) genotypes derive net benefit over harm from low-dose aspirin use, in the primary prevention setting ^8^.

Genetic variation is known to contribute to differences in drug response, as well as biological processes related to aspirin including lipid metabolism, platelet function, and vascular integrity ^9-12^. Polygenic scores (PGSs), which aggregate the effects of many different common genetic variants associated with a given trait or disease, can predict complex cardiovascular and metabolic responses ^13^, and may capture biological variation relevant to aspirin-induced bleeding. Some evidence suggests that genetic predisposition can modify aspirin’s effect on cardiovascular outcomes ^14,15^, however, the role of PGSs in predicting aspirin-related bleeding events remains largely unexplored.

In this post hoc genetic analysis of the ASPREE trial, we systematically screened 572 PGSs related to cardiovascular, coagulation, and hematologic traits to evaluate interactions with randomized low-dose aspirin allocation and incident major bleeding events. We hypothesized that there would be a polygenic score that interacted with aspirin use, and could identify individuals at increased bleeding risk. We also sought to explore the interaction of polygenic scores with aspirin for reducing major adverse cardiovascular events (MACE). Our objective was to identify genetic predictors that could influence both bleeding and cardiovascular risk on aspirin in the primary prevention setting, offering a potential future route to individualized preventive therapy.

## METHODS

### Study Design and Population

We conducted a post hoc genetic analysis of participants in the ASPREE trial, a randomized, placebo-controlled study of low-dose aspirin (100 mg of enteric-coated aspirin or matching placebo) in healthy older adults ^16^. Participants were aged ≥70 years from Australia and the United States (≥65 years for U.S. minorities), with no prior cardiovascular disease, dementia, physical disability, or chronic illness likely to limit survival to <5 years. Recruitment was conducted between March 2010 and December 2014. All participants provided written informed consent. The trial was approved by institutional review boards at all sites and registered at ClinicalTrials.gov (NCT01038583).

### End Points

The primary outcome was major bleeding, defined as a composite of hemorrhagic stroke, symptomatic intracranial bleeding, or clinically significant extracranial bleeding leading to transfusion, hospitalization, surgery, or death. Secondary outcomes included gastrointestinal bleeding, intracranial bleeding, and MACE (ischemic stroke, myocardial infarction, cardiovascular death, or other adjudicated coronary events). Outcomes were adjudicated by a committee of clinical experts blinded to treatment allocation.

### Genotyping and Polygenic Scores

Baseline DNA was genotyped using the Axiom 2.0 Precision Medicine Diversity Array. Standard quality control steps were applied ^17,18^, and imputation was performed using the TOPMed reference panel GRCh38 (hg38) ^19-21^. PGSs were obtained from the PGS Catalog for five trait categories: cardiovascular disease, cardiovascular measurement, hematologic measurement, inflammatory measurement, and lipid/lipoprotein measurement available up until the 22^nd^ of September, 2025 ^22,23^. PGSs were generated using PLINK 1.9 ^24^, and scores with low quality control metrics were excluded from the analysis. These included scores with low SNP availability (high data missingness), scores which did not use samples of European ancestry in genome wide association studies, and scores with detected issues related to distributional quality, whereby the approximate PGS distribution deviated substantially from a normal distribution (for details see Supplementary File).

### Statistical and Exploratory Analysis

For each PGS, Cox proportional-hazards models independently assessed interactions with aspirin allocation and major bleeding events, adjusted for age, sex, and the first 10 genetic principal components. P-values for interactions were Bonferroni-corrected. Significant PGSs were further analyzed with additional adjustment for baseline smoking, alcohol use, body mass index (BMI), hypertension, diabetes, chronic kidney disease (CKD), and prior aspirin or anti-inflammatory use. Distribution based quintile-specific analyses compared aspirin versus placebo effects for major bleeding, MACE, and bleeding subtypes. Baseline characteristics were compared across PGS quintiles using chi-square and ANOVA. Analyses were performed in R version 4.5.1. Exploratory analysis was performed based on results of polygenic score screening.

## RESULTS

### Polygenic Score Screening

Of 1,344 polygenic scores (PGSs) derived from 101 traits across five biologically relevant categories, 572 remained after quality control (Figure 1). Two triglyceride-related PGSs (PGS003144 and PGS003149) showed a significant interaction with aspirin allocation for major bleeding events after Bonferroni correction (adjusted P=0.034 and P=0.039, respectively). PGS003144, from the same source study as PGS003149, was selected for detailed follow-up analyses. PGS003144 was derived from a genome-wide association study of measured triglyceride levels in peripheral blood samples from 9,592 individuals ^25^.

**Figure 1.**
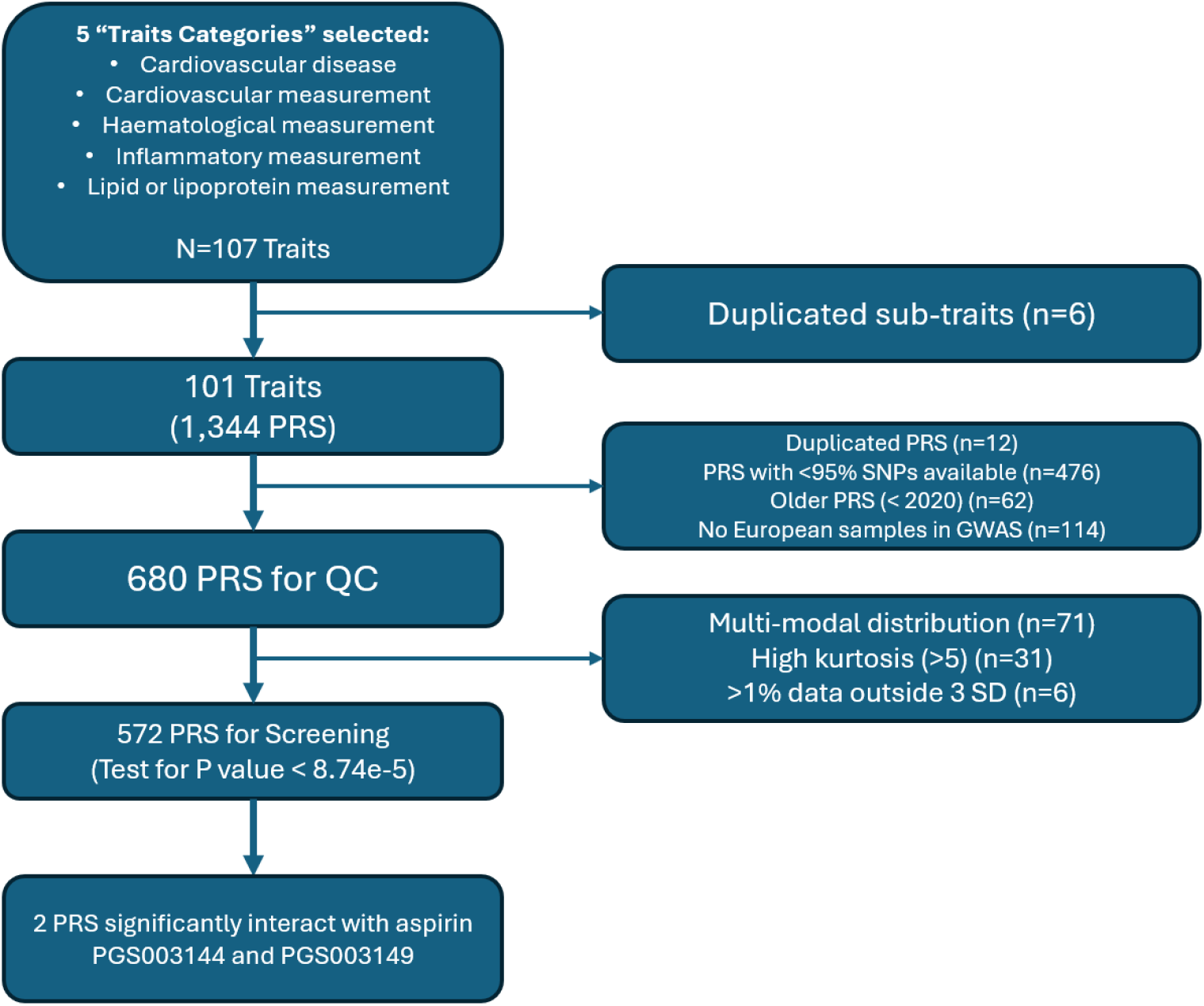
Flow diagram for polygenic score selection for subsequent screening.

### Study Population

A total of 13,571 genotyped ASPREE participants were included in the analysis. The mean age was 75 years (SD 4.3) and 54.8% were female. Baseline characteristics of the participants, stratified by PGS003144 quintiles are presented in Table 1. Overall, 6,758 participants (49.8%) received aspirin and 6,813 (50.2%) received placebo, with median follow-up of 4.6 years (IQR). Baseline differences across PGS quintiles included BMI (P = 0.011), hypertension (P = 0.005), diabetes (P < 0.0001), CKD (P = 0.004), alcohol use (P < 0.001), lipid levels (all P < 0.001), and statin use (P < 0.001).

**Table 1.** Baseline characteristics and trail period endpoints across quintiles.

| Baseline characteristics | All participants<br>(No. = 13571) | Quintile 1<br>(No. =2715) | Quintiles 2-4<br>(No. =8142) | Quintile 5<br>(No. =2714) | p-value |
| --- | --- | --- | --- | --- | --- |
| **Age, mean (SD)** | 75 (4.3) | 75.1 (4.4) | 75 (4.2) | 74.8 (4.2) | 0.053 |
| **Sex female, No. (%)** | 7441 (54.8) | 1473 (54.3) | 4462 (54.8) | 1506 (55.5) | 0.66 |
| **BMI, mean (SD)** | 28 (4.6) | 27.8 (4.6) | 28.1 (4.6) | 28.1 (4.5) | **0.011** |
| **Hypertension, No. (%)** | 9993 (73.6) | 1993 (73.4) | 6038 (74.2) | 2022 (74.5) | **0.005** |
| **Diabetes, No. (%)** | 1321 (9.7) | 194 (7.2) | 796 (9.8) | 331 (12.2) | **<0.0001** |
| **CKD, No. (%)** | 3256 (24) | 610 (22.5) | 1935 (23.8) | 711 (26.2) | **0.004** |
| **Smoking status, No. (%)** |  |  |  |  |  |
| Current/Former | 6022 (44.4) | 1193 (43.9) | 3599 (44.2) | 1230 (45.3) | 0.52 |
| Never | 7549 (55.6) | 1522 (56.1) | 4543 (55.8) | 1484 (54.7) |  |
| **Alcohol use, No. (%)** |  |  |  |  |  |
| Current | 10702 (78.9) | 2221 (81.8) | 6369 (78.2) | 2112 (77.8) | **0.0001** |
| Former/Never | 2869 (21.1) | 494 (18.2) | 1773 (21.8) | 602 (22.2) |  |
| **Lipids** |  |  |  |  |  |
| HDL mmol/L, median (IQR)<sup>1</sup> | 1.5 (0.5) | 1.6 (0.6) | 1.5 (0.5) | 1.4 (0.5) | **<0.0001** |
| LDL mmol/L, median (IQR)<sup>2</sup> | 3 (1.1) | 3.1 (1.1) | 3 (1.1) | 3 (1.4) | **<0.0001** |
| Triglycerides mmol/L, median (IQR)<sup>3</sup> | 1.2 (0.7) | 1 (0.6) | 1.2 (0.7) | 1.5 (0.9) | **<0.0001** |
| **Medication** |  |  |  |  |  |
| Statins, No. (%) | 4180 (30.8) | 636 (23.4) | 2472 (30.4) | 1072 (39.5) | **<0.0001** |
| NSAID, No. (%) | 1795 (13.2) | 336 (12.4) | 1093 (13.4) | 366 (13.5) | 0.34 |
| Previous aspirin use, No. (%) | 1300 (9.6) | 247 (9.1) | 789 (9.7) | 264 (9.7) | 0.63 |
| Trail aspirin allocation, No. (%) | 6758 (49.8) | 1342 (49.4) | 4020 (49.4) | 1396 (51.4) | 0.16 |
| **Endpoints, No. (%)** |  |  |  |  |  |
| Major Bleeding | 414 (3.1) | 87 (3.2) | 249 (3.1) | 78 (2.9) | 0.78 |
| MACE | 464 (3.4) | 80 (2.9) | 288 (3.5) | 96 (3.5) | 0.31 |
| Gastrointestinal Bleeding | 169 (1.3) | 35 (1.3) | 102 (1.3) | 32 (1.2) | 0.093 |
| Intracranial Bleeding | 112 (0.8) | 26 (0.96) | 70 (0.86) | 16 (0.59) | 0.28 |

BMI, body mass index; CKD, chronic kidney disease; HDL, high-density lipoprotein; LDL, low-density lipoprotein; MACE, major adverse cardiovascular events, NSAID, non-steroidal anti-inflammatory drug use.
<sup>1</sup> Data on 341 participants missing. <sup>2</sup> Data on 385 participants missing. <sup>3</sup> Data on 159 participants missing.

### Genetic Stratification

In the lowest quintile of the PGS003144 triglyceride PGS distribution (Q1), aspirin use versus placebo was associated with a markedly higher risk of major bleeding (HR 2.28; 95% CI, 1.45–3.58; P = 0.00036), including gastrointestinal bleeding (HR 3.17; 95% CI, 1.48–6.80; P = 0.0029), and intracranial bleeding (HR 4.10; 95% CI, 1.54–11.0; P = 0.0049) after adjusting for multiple covariates (Table 2). However, no association was observed in Q1 for MACE (HR 1.04; 95% CI, 0.67–1.62; P = 0.86) in individuals randomized to aspirin versus placebo.

**Table 2.**
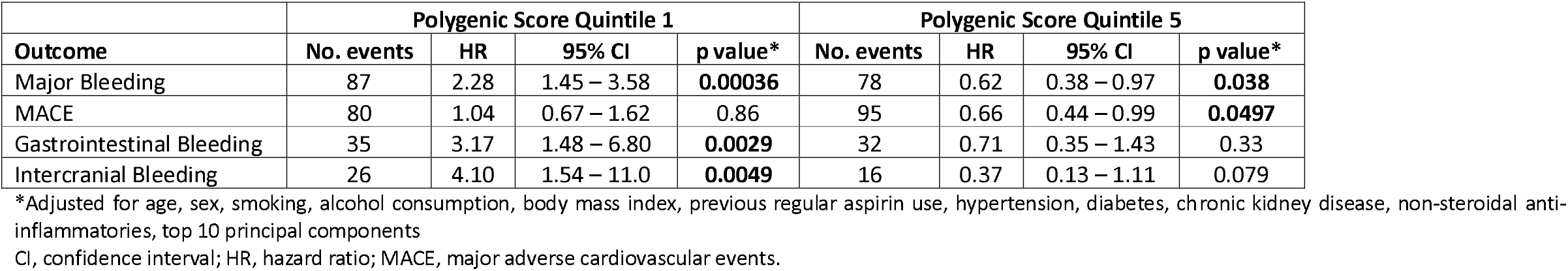
Outcome risk of aspirin vs placebo, within PGS003144 polygenic score quintile stratification, fully adjusted model.

In contrast, participants in the highest PGS quintile (Q5) randomized to aspirin versus placebo had lower risk of major bleeding (HR 0.62; 95% CI, 0.38–0.97; P = 0.038) and MACE (HR 0.66; 95% CI, 0.44–0.99; P = 0.0497), with no significant effect on gastrointestinal or intracranial bleeding. Six-year cumulative incidence of bleeding, stratified by aspirin and placebo allocation, for a) all participants, b) PGS quintile 1, and c) PGS quintile 5, can be seen in Figure 2 (MACE results in Supplementary Figure S1). All associations were similar after a simpler adjustment of age, sex, and top 10 genetic principal components (Supplementary Table S1).

**Figure 2.**
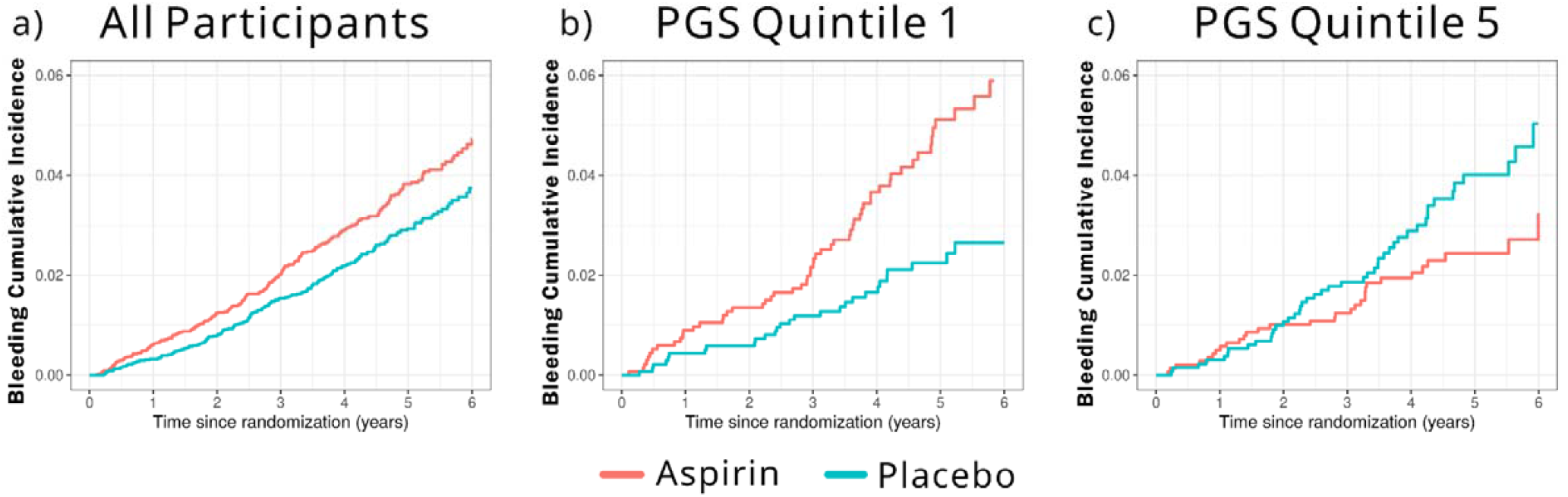
Aspirin use and 6-year cumulative incidence for major bleeding in all participants and within PGS003144 genetic quintiles. **a)** All participants: total aspirin group = 5831, MH in aspirin group = 227; total placebo group = 5907, MH in placebo group = 178 **b)** Triglyceride genetic quintile 1: total aspirin group = 1149, MH in aspirin group = 57; total placebo group = 1193, MH in placebo group = 28 **c)** Triglyceride genetic quintile 5: total aspirin group = 1213, MH in aspirin group = 31; total placebo group = 1158, MH in placebo group = 45 PGS; Polygenic Score.

### Exploratory Analyses

Baseline fasting triglyceride levels were available for all participants of the ASPREE trial. Upon exploratory analysis of these measured triglyceride levels, we found moderate correlation between measured levels and the PGS003144 triglyceride PGS (r = 0.30; P < 0.0001). Participants in the lowest measured triglyceride quintile (Q1) and assigned to aspirin had a higher risk of major bleeding (HR 1.68; 95% CI, 1.12–2.51; P = 0.01) and intracranial bleeding (HR 3.10; 95% CI, 1.39–6.91; P = 0.006) when compared to placebo. No differences were observed in the other measured triglyceride level quintiles (Table 3).

**Table 3.**
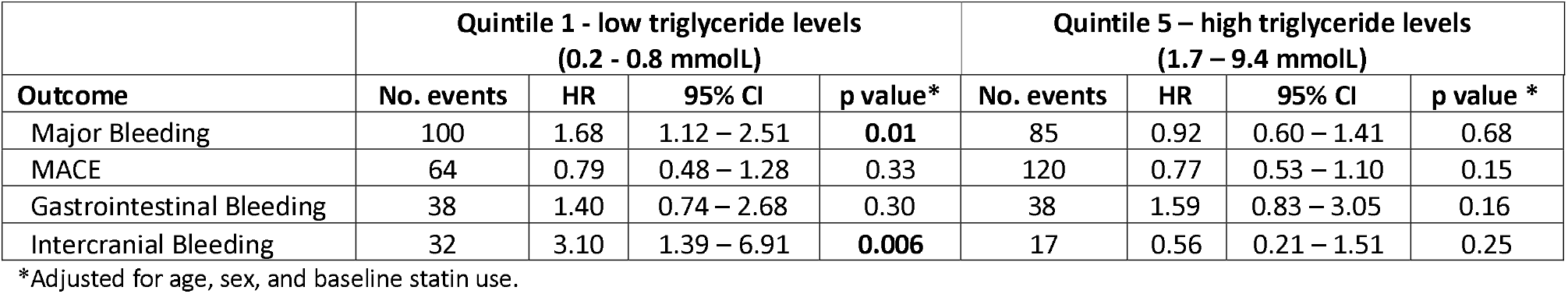
Aspirin vs placebo stratified by triglyceride level quintiles.

## DISUSSION

In this post hoc genetic analysis of 13,571 participants from the ASPREE randomized control trial, a triglyceride-level PGS identified differential responses to aspirin for major bleeding and MACE. Participants in the lowest quintile of the triglyceride PGS had increased risk of major bleeding, gastrointestinal and intracranial bleeding with aspirin, whereas those in the highest quintile experienced lower major bleeding risk and a 1.5-fold reduction in MACE. These findings suggest that genetic variation in triglyceride-related pathways (and in measured triglyceride levels), can identify differential bleeding and cardiovascular risk with aspirin, in the primary prevention setting.

Our findings add a new dimension to the ongoing debate over aspirin use for primary prevention of cardiovascular disease in older adults. Although aspirin has long been recognized for its cardioprotective effects, recent evidence has suggested that the harms from bleeding outweigh any vascular benefit in primary prevention ^3,5,6^. This has resulted in the U.S. Preventive Services Task Force (USPSTF) recommending against routine aspirin use for primary prevention in adults aged 60 years or older ^26^. However, the above trials reflect an overall harm-to-benefit ratio calculated in all trial participants, without stratification or subgrouping. The USPSTF population-level guidance does not account for potential interindividual variation (e.g. genetic) in aspirin response.

Our results suggest that genetic factors (related to triglyceride levels) may identify subgroups for whom the net benefit:harm ratio shifts favorably, potentially enabling a more individualized approach to aspirin therapy in the primary prevention setting, and a reduction in the harms of bleeding. Previously studies have identified possible subgroups with differential bleeding or cardiovascular risks on aspirin, based on analysis of traditional clinical risk factors rather than genetic ^7^. Our study now adds to this, specifically investigating the role of genetics in the aspirin response. Further, our previous study also identified different genetic factors (specific lipoprotein(a) genotypes)interacting with aspirin for MACE reduction. We now report genetic variation related to triglyceride levels that appears to identify differential harm and bleeding risk. Combining previous findings with our current data suggests that lipid-related genotypes and lipid metabolism have a strong influence in differential aspirin response, both with regards to cardiovascular benefits and bleeding harms. These findings raise the possibility of future strategies where genetically-guided or personalized aspirin therapy may be possible.

Triglycerides play a central role in the transport and storage of fatty acids in plasma and tissues ^27^. They are a key component of the lipid profile and have been primarily studied in the context of cardiovascular risk or malnutrition, with elevated levels linked to atherosclerosis and ischemic events ^28^. Fewer studies, however, have examined the relationship between low triglyceride levels and hemorrhagic outcomes. Epidemiologic evidence indicates that low triglyceride levels (or hypotriglyceridemia) (<50 mg/dL) are associated with increased risk of hemorrhagic stroke, particularly among men, individuals with hypertension, and those with low cholesterol levels ^29^. Similarly, low measured triglyceride levels have been linked to intracerebral hemorrhage and deep or infratentorial cerebral microbleeds, independent of high-density lipoprotein or low-density lipoprotein cholesterol levels ^30^. However, low measured triglyceride levels in older people may typically reflect age related changes, such as frailty ^31,32^, or related malnutrition. A low polygenic score for triglycerides, by contrast, reflects an underlying genetic propensity from birth, toward lower triglyceride levels. Our findings suggest that low polygenic triglycerides, or related metabolic pathways, may contribute to vascular integrity and hemostatic balance with regards to the aspirin response. To our knowledge, this association and relationship has not been reported previously.

Conversely, hypertriglyceridemia has been associated with enhanced platelet reactivity and reduced responsiveness to aspirin in patients with ischemic stroke or diabetes ^33,34^. This pattern aligns with our observation that genetically higher triglyceride levels were associated with lower bleeding risk but also with reduced MACE on aspirin, implying a complex relationship between triglyceride metabolism, vascular stability, and aspirin pharmacodynamics ^35-37^. The biological mechanisms underlying this association remain to be clarified but may involve membrane lipid composition, platelet activation pathways, or endothelial resilience.

Our exploratory analysis of measured triglyceride levels provided complementary evidence of our genetic findings: participants with low measured triglyceride levels assigned to aspirin had a higher risk of major and intracranial bleeding, whereas those with high measured triglyceride levels did not. The moderate correlation between measured and genetically predicted triglycerides supports a potential shared pathway, but also indicates that the genetic score may capture different and broader underlying lipid-related biology, rather than environmentally or dietary-influenced plasma triglyceride levels.

### Strengths and Limitations

This study has several strengths, including the large, well-characterized, randomized trial population; systematic screening of polygenic scores; rigorous ascertainment and adjudication of outcomes by expert committees masked to randomized treatment allocation; and comprehensive genetic data enabling polygenic screening. Nonetheless, limitations should be acknowledged. The analysis was post hoc and hypothesis-generating, and replication in independent cohorts is needed. The ASPREE cohort was predominantly of European ancestry, which may limit generalizability. In addition, while polygenic scores capture cumulative genetic risk from common alleles, they do not directly identify rare variants, causal variants or explanatory genetic mechanisms.

## Conclusions

A triglyceride-related PGS modifies aspirin-associated bleeding and MACE risk in older adults, and can effectively identify individuals at increased future bleeding risk. These results provide evidence that genetic profiling could enable a future pathway towards individualized aspirin (or other drug) therapy in primary prevention, potentially optimizing cardiovascular benefit while minimizing harm. Further investigation in more diverse additional populations and randomized trials is warranted to validate these findings.

## Data Availability

Due to the nature of genomics, the de-identified data we analyzed are not publicly available, however all data produced in the present study are available upon reasonable request to the authors.

## Data sharing statement

Due to the nature of genomics, the de-identified data we analyzed are not publicly available, but requests to the corresponding author for the data will be considered on a case-by-case basis.

## Acknowledgements

We thank the ASPREE (Aspirin in Reducing Events in the Elderly) trial staff and participants, and the general practitioners and staff of the medical clinics who cared for the participants.

## Authors’ contributions

PDF, PL, and CY conceptualized the study. PDF undertook data analysis, interpretation, and writing of the first draft manuscript with the input of PL, CY and JM. All authors including PDF, CY, CT, SMH, CAB, MRN, AMT JJM and PL contributed to the writing, direction and editing of the final draft. All authors read and approved the final manuscript.

