## Supplementary File 1 for "Triglyceride Polygenic Score Identifies Differential Bleeding and Cardiovascular Risk with Aspirin in Primary Prevention"

**Contents**

**Page 1:** Detailed Methods
**Page 4:** Supplementary Figure S1

**Page 5:** Supplementary Table S1

**Page 6:** References

**Detailed Methods**

**Genotyping and Polygenic Scores**

Genotyping was carried out on baseline DNA using the Axiom 2.0 Precision Medicine Diversity Array (Thermo Fisher Scientific, CA). Variant calling of ~850 000 variants was done using a customised pipeline, aligning data to the human reference genome GRCh38 (hg38). Low-quality samples and single nucleotide polymorphisms (SNPs) were removed during quality control steps (1). Genetic ancestry and relationships were inferred (2) and imputation was performed using the TOPMed Imputation Server with the TOPMed ‘r3’ reference panel, containing 133,597 genetically diverse reference samples with data from over 445 million genetic variants (3-5). Variants were removed in post-imputation QC if imputation quality score R2 was <0.3.

We downloaded polygenic scores (PGSs) under 5 trait categories from the PGS Catalog (6, 7) that may be associated with aspirin and bleeding risk, including the categories of ‘Cardiovascular disease’, ‘Cardiovascular measurement’, ‘Haematological measurement’, ‘Inflammatory measurement’, and ‘Lipid or lipoprotein measurement’ available up until the 22^nd^ of September 2025. PGSs were generated in our data using PLINK 1.9 (8). Duplicate traits between categories and duplicate PGSs between traits were removed. PGSs that did not have >95% SNPs available in our dataset, or PGSs that did not use European samples in their genomic wide association studies (GWAS), or PGSs published prior to 2021 were removed. Based on central limit theorem for PGSs, further quality control was carried out to assure approximate normal distribution (9). Silverman modality test identified PGSs with multimodal distribution and were removed (10). Further, PGSs with high kurtosis (long tails in distribution i.e. >5) and PGSs with over 1% of their data outside of 3 standard deviations were also removed.

**Statistical and Exploratory Analysis**

For each PGS, Cox proportional-hazards models were fitted with major hemorrhage as the primary outcome, including terms for aspirin allocation, PGS, and their interaction, adjusted for age, sex, and the first 10 principal components. Interaction P values were Bonferroni-corrected for multiple testing. PGSs showing significant interactions were further evaluated in models adjusted for baseline smoking status, alcohol consumption, body-mass index, hypertension, diabetes, chronic kidney disease, and prior use of aspirin or anti-inflammatory drugs. Biological samples were taken at baseline, including fasting high-density lipoprotein, low-density lipoprotein and triglycerides (mmol/L). Baseline statin use in this study included any use of atorvastatin, rosuvastatin, simvastatin, or pravastatin. Baseline characteristics were compared between PGS quintile groups using chi-square and ANOVA. PGS quintiles were examined separately, comparing aspirin versus placebo effects in the lowest (Q1) and highest (Q5) quintiles for major hemorrhage, MACE, gastrointestinal and intercranial bleeding. All analyses were conducted in R version 4.5.1.

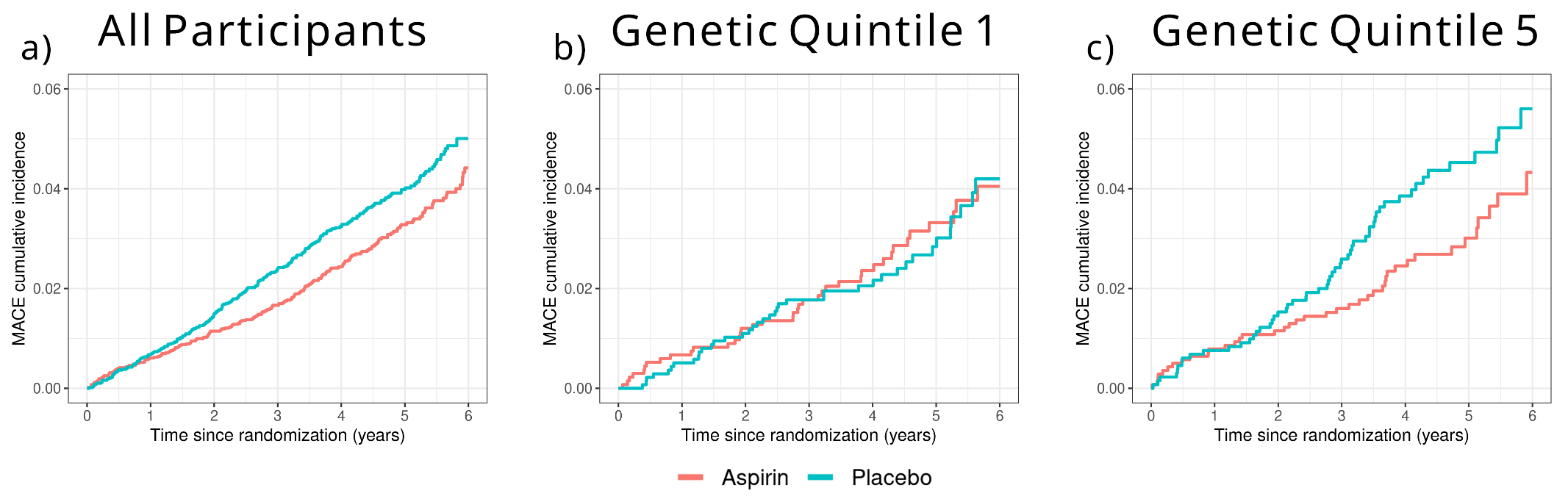

**Figure S1. Aspirin use and 6-year cumulative incidence for MACE in all participants and within** **PGS003144 genetic quintiles.**

**a)** All participants: total aspirin group = 5829, MACE in aspirin group = 201; total placebo group = 5908, MACE in placebo group = 253

**b)** Triglyceride genetic quintile 1: total aspirin group = 1151, MACE in aspirin group = 39; total placebo group = 1198, MACE in placebo group = 39

**c)** Triglyceride genetic quintile 5: total aspirin group = 1213, MACE in aspirin group = 40; total placebo group = 1154, MACE in placebo group = 55

**Table S1. Outcome risk of aspirin vs placebo, within PGS003144 quintile stratification (Model 1*).**

|  | **PGS Quintile 1** | | | | **PGS Quintile 5** | | | |
| --- | --- | --- | --- | --- | --- | --- | --- | --- |
| **Outcome** | **n events** | **HR** | **95% CI** | **p-value** | **n events** | **HR** | **95% CI** | **p-value** |
| Major Hemorrhage | 87 | 2.24 | 1.42 - 3.51 | **0.00048** | 78 | 0.62 | 0.40 – 0.98 | **0.0406** |
| MACE | 80 | 1.02 | 0.66 - 1.59 | 0.92 | 96 | 0.65 | 0.44 – 0.98 | **0.0404** |
| Gastrointestinal Bleeding | 35 | 3.10 | 1.44 - 6.06 | **0.0037** | 32 | 0.70 | 0.35 – 1.42 | 0.32 |
| Intercranial bleeding | 26 | 4.14 | 1.55 – 11.0 | **0.0044** | 16 | 0.38 | 0.13 – 1.12 | 0.08 |

MACE, major adverse cardiovascular events, *Adjusted for age, sex and top 10 PCs
